# Impact of COVID-19 on diagnoses, monitoring and mortality in people with type 2 diabetes: a UK-wide cohort study involving 14 million people in primary care

**DOI:** 10.1101/2020.10.25.20200675

**Authors:** Matthew J. Carr, Alison K. Wright, Lalantha Leelarathna, Hood Thabit, Nicola Milne, Naresh Kanumilli, Darren M. Ashcroft, Martin K. Rutter

## Abstract

**AIMS:** To compare trends in diagnoses, monitoring and mortality in patients with type 2 diabetes, before and after the first COVID-19 peak.

**METHODS:** We constructed a cohort of 25 million patients using electronic health records from 1831 UK general practices registered with the Clinical Practice Research Datalink (CPRD), including 14 million patients followed between March and December 2020. We compared trends using regression models and 10-year historical data. We extrapolated the number of missed/delayed diagnoses using UK Office for National Statistics data.

**RESULTS:** In England, rates of new type 2 diabetes diagnoses were reduced by 70% (95% CI 68%-71%) in April 2020, with similar reductions in Northern Ireland, Scotland and Wales. Between March and December, we estimate that there were approximately 60,000 missed/delayed diagnoses across the UK. In April, rates of HbA_1c_ testing were greatly reduced in England (reduction: 77% (95% CI 76%-78%)) with more marked reductions in the other UK nations (83% (83-84%)). Reduced rates of diagnosing and monitoring were particularly evident in older people, in males, and in those from deprived areas. In April, the mortality rate in England was more than 2-fold higher (112%) compared to prior trends, but was only 65% higher in Northern Ireland, Scotland and Wales.

**CONCLUSIONS:** As engagement increases, healthcare services will need to manage the backlog and anticipate greater deterioration of glucose control due to delayed diagnoses and reduced monitoring in those with pre-existing diabetes. Older people, men, and those from deprived backgrounds will be groups to target for early intervention.

**RESEARCH IN CONTEXT:** *What is already known about this subject?:*

- The higher COVID-related death rate in people with diabetes has been well-documented
- A study involving the residents of Salford, UK showed 135 fewer diagnoses of type 2 diabetes than expected between March and May 2020, which amounted to a 49% reduction in activity
- There is limited data on the impact of the COVID-19 pandemic on the diagnosis and monitoring of type 2 diabetes

*What is the key question?:*

- What has been the impact of the COVID-19 pandemic on the diagnosis and monitoring of type 2 diabetes across the UK?

*What are the new findings?:*

- Across the UK, the rate of new type 2 diabetes diagnoses was reduced by up to 70% in April 2020 compared to 10-year historical trends
- Between March and December 2020, it is estimated that 60,000 people have had a missed or delayed diagnosis
- The frequency of HbA_1c_ monitoring in type 2 diabetes was reduced by 77-83% in April 2020 and by 31-37% overall between March and December 2020

*How might this impact on clinical practice in the foreseeable future?:*

- During this pandemic and associated lockdowns, effective public communications should ensure that patients remain engaged with diabetes services including HbA_1c_ screening and monitoring

## INTRODUCTION

The COVID-19 pandemic has had major health and economic effects across the world. So far in the UK, there have been more than 100,000 COVID-related deaths with disproportionate impacts in people with diabetes; nearly a third of all COVID-related deaths having occurred in people with diabetes ^1–3^.

The impact on the NHS, and in particular on diabetes services, has been enormous, with the suspension of much routine care. As the COVID-19 pandemic continues, there is an urgent need to minimise the harm done through suspension of routine services and to prioritise care and resources to areas of greatest need.

The diagnosis of type 2 diabetes occurs almost exclusively in primary care ^4^. Earlier detection of type 2 diabetes and treatment are critically important as increased exposure to hyperglycaemia is associated with increased risk of long-term complications including cardiovascular disease and mortality ^5^. Therefore, any delays in normal screening and testing processes for type 2 diabetes as a result of lower general practice attendance due to COVID-19 will have an impact on future diabetes-related complications.

There is limited data on the indirect consequences of the COVID-19 pandemic on the incidence and monitoring of diabetes in primary care. Likewise, there is limited information on COVID-19 impacts on mortality rates in people with diabetes during and after the first wave of COVID-19.

We used a large primary care longitudinal dataset, broadly representative of the UK population, aiming to compare: i) the UK-wide incidence of type 2 diabetes; ii) the frequency of HbA_1c_ testing; and iii) mortality rates in people with type 2 diabetes, before and after the first nationwide COVID-19 lockdown in March 2020. We compared observed and predicted rates using data covering ten years prior to the pandemic.

Since older people and more socially disadvantaged groups have been disproportionally affected by COVID-19 infections, and since the same groups may be more adversely impacted by the unintended consequences of government interventions, we aimed to study variation in outcomes by gender, age group, deprivation level and region.

## METHODS

### Data sources

We conducted a retrospective cohort study using primary care electronic health records obtained from the Clinical Practice Research Datalink (CPRD) *Aurum* and *GOLD* databases ^6,7^. The study population consisted of 21,797,864 patients from 1470 general practices in England, with a further 40 practices in Northern Ireland (376,143 patients), 206 practices in Scotland (1,930,762 patients), and 112 in Wales (1,319,053 patients).

A total of 24,440,354 patients were included for estimation of the expected rates in the pre-COVID-19 period (January 2010 to February 2020). The CPRD contains anonymised consultation records and includes patient demographic information, symptoms, diagnoses, medication prescriptions, and date of death. We also examined practice-level Index of Multiple Deprivation (IMD) quintiles ^8^, a measure representing an area’s relative level of deprivation, ranked within each UK nation.

### Definitions, measurements and clinical coding

To enable comparisons of rates before and after the start of the COVID-19 outbreak, we included patient records from January 2010 to establish long-term trends and patterns of seasonality. We focussed primarily on reporting observed versus expected rates from 1^st^ March 2020 to 10^th^ December 2020. First, we estimated incidence rates of type 2 diabetes diagnoses, new prescriptions for metformin (the most commonly prescribed medication in new-onset type 2 diabetes) and insulin, and rates of HbA_1c_ testing and mortality in people with type 2 diabetes.

Incident type 2 diabetes was identified from Read/SNOMED/EMIS codes used in CPRD *GOLD* and *Aurum* (see https://clinicalcodes.rss.mhs.man.ac.uk). In line with guidance from the CPRD’s central administration, data from the *Aurum* and *GOLD* databases were analysed separately, with data from *Aurum* restricted to English practices and *GOLD* providing information on practices in Northern Ireland, Scotland and Wales. The use of two discrete data sources also enabled independent replication of our findings. All code lists and medication lists were verified by two senior clinical academics (a diabetologist: MKR, and a senior academic pharmacist: DMA).

### Study design

For each patient, we defined a ‘period of eligibility’ for study inclusion which commenced on the latest of: the study start date (1st January 2010); the patient’s most recent registration with their practice; the date on which data from the practice was deemed to be ‘up-to-standard’ by the CPRD. A patient’s period of eligibility ended on the earliest of: registration termination; the end of data collection from their practice; death. For incident diagnoses and prescriptions, we also applied a ‘look-back’ period during which a patient was required to have been registered for at least a year prior to the event. Flow diagrams illustrating the delineation of the study cohorts using CPRD *Aurum* and *GOLD* are presented in **Supplementary Figures S1 and S2** respectively. The denominator for the incidence rates was the aggregate person-months at risk for the whole eligible study population. Mortality and testing rates in people with type 2 diabetes were calculated using the person-months at risk from all those with type 2 diabetes as the denominator. Incidence, mortality and testing rates were stratified by gender, age group (<18, 18-29, 30-44, 45-64, 65-79 and ≥80 years), practice-level deprivation (IMD quintiles) and region (in England) or nation (in the rest of the UK).

### Statistical Analysis

The data were structured in a time-series format with event counts and ‘person-months at risk’ aggregated (by year and month) with stratification by gender, age group, deprivation quintile and region (or nation in *GOLD*). Mean-dispersion negative binomial regression models were used to estimate expected monthly event counts from March 2020 onward based on antecedent trends since 2010. The natural logarithm of the denominator (person-months at risk) was used as an offset in each regression model. To account for possible seasonality and long-term linear trends, calendar month was fitted as a categorical variable and time as a continuous variable with the number of months since the start of the study serving as the unit of measurement. For each month studied, observed and expected event counts were converted to rates using the observed person-month denominator. The monthly expected rates, and their 95% confidence intervals, were plotted against the observed rates. As they share a common denominator, differences between expected and observed monthly rates are expressed as a percentage ‘rate reduction (or increase)’.

Extrapolated estimates of the number of missed (or delayed) diagnoses of type 2 diabetes were derived using the discrepancy between observed and expected frequencies from March 2020 onward, and approximations of the proportional representation of the populations of England and the rest of the UK (in CPRD *Aurum* and *GOLD* respectively) using data from the Office for National Statistics ^9^.

All data processing and statistical analyses were conducted using Stata version 16 (StataCorp LP, College Station, TX). We followed RECORD (REporting of studies Conducted using Observational Routinely-collected health Data) guidance ^10^.

## RESULTS

### Study cohort

Our focus was on the impact of the COVID-19 pandemic between March and December 2020. Using the inclusion criteria described in the Study Design, a mixed cohort was utilised consisting of patients whose period of eligibility began before 1st March 2020 and those who became eligible for inclusion between 1st March 2020 and 10th December 2020. The study cohort was comprised of 14,929,251 patients (median (IQR) age: 41 (25, 59) years, 50% female) of whom 790,377 had type 2 diabetes. Of those with type 2 diabetes, the median (IQR) age was 67 (57, 76) years, 44% were female and 25% lived in an area that was in the most deprived quintile compared to the rest of the UK.

### Impacts of COVID-19 on diagnosis, prescribing and HbA_1c_ monitoring in England

In April 2020, the rate of new diagnoses of type 2 diabetes in English primary care was reduced by 70% (95% CI 68% to 71%) compared to the expected rates based on 10-year historical trends (**Figure 1A; Supplementary Table S1**). Prior to March 2020, rates of type 2 diabetes diagnoses in English practices were higher in older individuals, in men, and in people from deprived areas. These groups experienced the greatest reductions in rates for new type 2 diabetes diagnosis at the time of the first COVID-19 peak (**Supplementary Figure S3)**. The reduced rates of type 2 diabetes diagnosis in April 2020 were mirrored by reduced rates of new metformin prescriptions in English practices (reduction: 53% (95% CI 51% to 55%; **Figure 1B; Supplementary Table S1**). In April, rates of HbA_1c_ testing in England were greatly reduced in people with type 2 diabetes (reduction: 77% (95% CI: 76% to 78%)); **Figure 1C; Supplementary Table S1**; with the largest reductions observed in older patients (**Supplementary Figure S4A**). Insulin prescribing was reduced by 27% (24% to 31%); (**Figure 1D; Supplementary Table S1**). Reductions in rates of new prescribing for both metformin and insulin were most evident in people aged over 65 years (metformin **Supplementary Figure S5A;** insulin**: Supplementary Figure S6A**).

**Figure 1.**
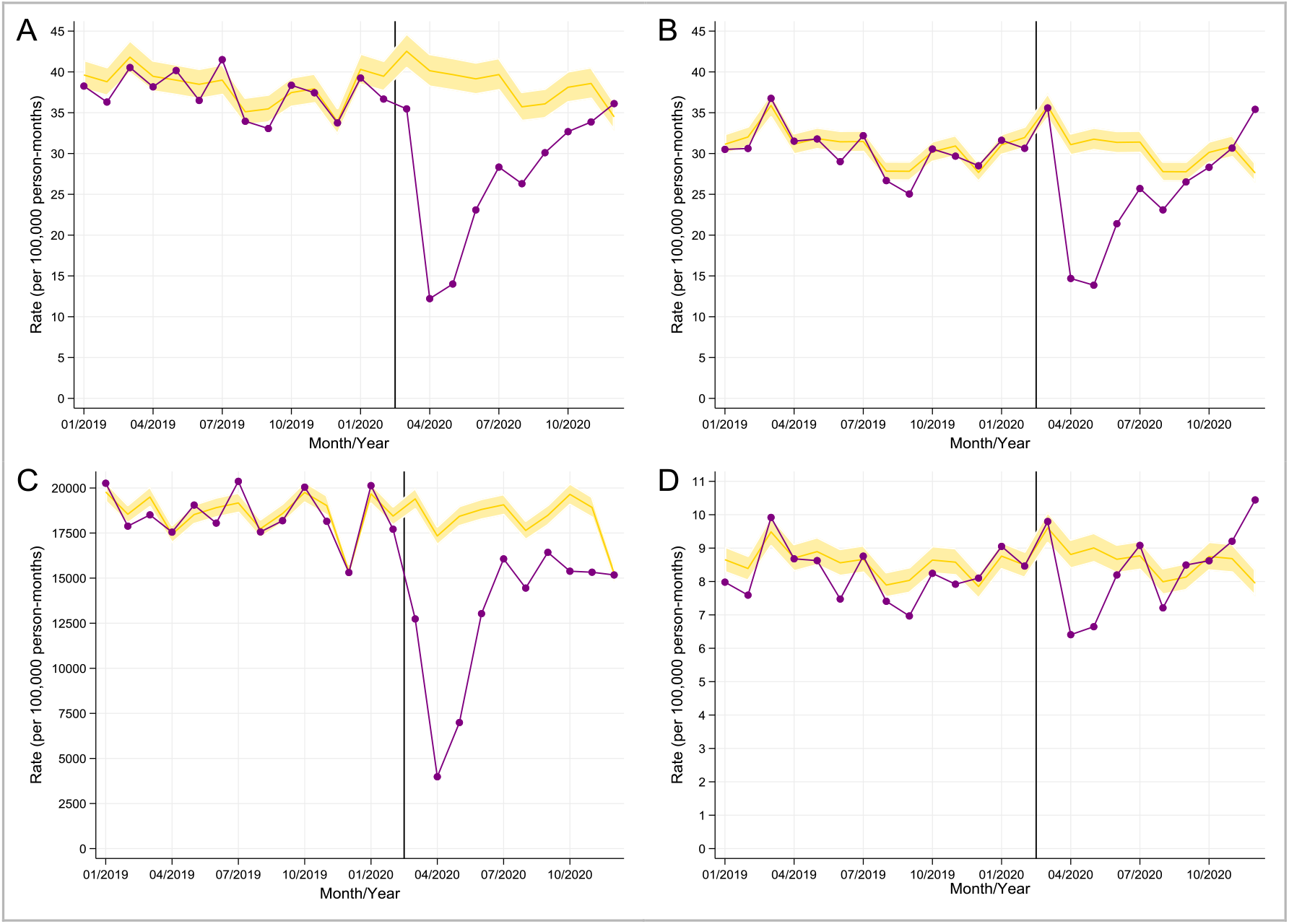
Comparison of observed (purple line) and expected (gold line with shaded area representing 95% CI) monthly rates in primary care before and after the start of the COVID-19 pandemic in England. *A*: incident diagnoses of type 2 diabetes, *B*: new metformin prescriptions, *C*: HbA_1c_ monitoring in patients with type 2 diabetes, *D*: new insulin prescriptions. Rates were derived using data from CPRD *Aurum* covering 21,797,864 patients. X-axis markers are mid-month. The vertical line denotes 1^st^ March 2020.

The reduced rates of diagnosis, new insulin/metformin prescribing and HbA_1c_ testing increased gradually between May and December 2020, though levels remained well below expected rates based on 10-year historical data for the majority of the period (**Figure 1A-D**). Overall in English practices, between 1^st^ March and 10^th^ December 2020, the rate of diagnosis of type 2 diabetes was reduced by 32% (95% CI: 28% to 35%), metformin prescribing was reduced by 20% (16% to 23%), insulin prescribing fell by 5% (0.2% to 10%), and HbA_1c_ testing in people with type 2 diabetes was reduced by 31% (29% to 33%); **Supplementary Table S1**.

### Impact of COVID-19 on mortality in England

In April 2020, mortality rates in people with type 2 diabetes in England were more than 2-fold higher compared to prior trends (mortality rate increase: 112% (95% CI: 104% to 120%); **Figure 2A; Supplementary Table S1**). Peaks in mortality were seen particularly in individuals aged over 65 years (**Supplementary Figure S7A**). Mortality rates returned to expected levels in people with type 2 diabetes and sub-groups between June and September 2020 (**Figure 2A**). Overall, between 1^st^ March and 10^th^ December 2020, the rate of mortality in people with type 2 diabetes in English practices was increased by 19% (95% CI: 14% to 23%); **Supplementary Table S1**.

**Figure 2.**
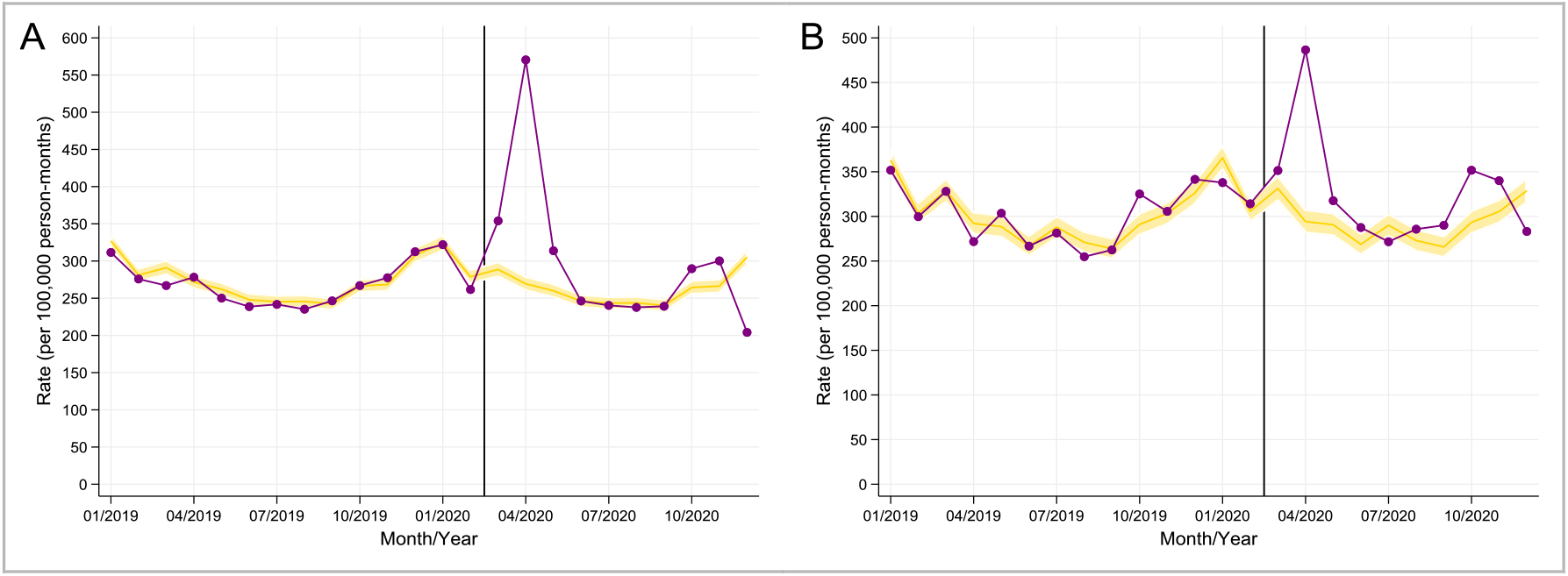
Comparison of observed (purple line) and expected (gold line with shaded area representing 95% CI) monthly mortality rates in people with type 2 diabetes before and after the start of the COVID-19 pandemic, in England (*A*) and in Northern Ireland, Scotland and Wales (*B*). Rates were derived using data from CPRD *Aurum* covering 21,797,864 patients in England, and from *CPRD GOLD* covering 3,625,958 patients. X-axis markers are mid-month. The vertical line denotes 1^st^ March 2020.

### Impacts of COVID-19 in Northern Ireland, Scotland and Wales (CPRD GOLD)

The temporal trends noted above in England (CPRD *Aurum* practices) were similar overall in Northern Ireland, Scotland and Wales (CPRD *GOLD* practices) with some notable exceptions. In April 2020, percentage reductions in the incidence of type 2 diabetes, metformin and insulin prescribing in Northern Ireland, Scotland and Wales were similar to reductions in England (**Supplementary Figure S8; Supplementary Table S2**). However, in Northern Ireland, Scotland and Wales (CPRD *GOLD*), the reduction in the rate of HbA_1c_ testing in people with type 2 diabetes was greater (*GOLD* vs. *Aurum*: 83% vs. 77%) and the increase in mortality rate was smaller (*GOLD* vs. *Aurum*: 65% vs. 112%; **Supplementary Tables S1 and S2**).

During the nine months between 1^st^ March and 10^th^ December 2020, there were smaller percentage reductions in incident type 2 diabetes and new metformin prescribing in practices based in Northern Ireland, Scotland and Wales (CPRD *GOLD*) compared to in England (*GOLD* vs. *Aurum*: incident type 2 diabetes reduced: 21% vs. 32%; metformin prescribing reduced: 12% vs. 20%; **Supplementary Tables S1 and S2**). Over the same nine month period, the overall reduction in HbA_1c_ testing in type 2 diabetes was greater in CPRD *GOLD* practices based in Northern Ireland, Scotland and Wales (*GOLD* vs. *Aurum*: 37% vs. 31%) but the mortality rate increase was lower than in England (*GOLD* vs. *Aurum*: 13% vs. 19%; **Figure 2; Supplementary Tables S1 and S2)**.

## DISCUSSION

Using primary care data from more than 14 million people in the UK, and 10-year historical data, we have shown that following the first nationwide ‘lockdown’, the indirect consequences of the COVID-19 pandemic led to: i) a 68-70% reduction in new diagnoses of type 2 diabetes in April 2020, with older individuals, males, and people from deprived areas experiencing the greatest reduction in diagnosis rates; ii) a 77-83% reduction in HbA_1c_ testing; iii) a reduction in metformin and insulin prescribing, particularly in older people with type 2 diabetes, supporting the reduced rates of diagnosis and monitoring; and iv) a short-term 112% increase in mortality rate in people with type 2 diabetes in England and a 65% increased mortality rate across the rest of the UK. There is limited prior data on the impact of the COVID-19 pandemic on the diagnosis of type 2 diabetes. A study using primary care data from Salford, UK showed 135 fewer diagnoses of type 2 diabetes than expected between March and May 2020, which amounted to a 49% reduction in activity ^11^. Here we extend these observations by assessing primary care data across the UK over a longer period of time and by providing supplementary data on HbA_1c_ testing and mortality. We show that the reduced rate of diagnosis applies to all areas of the UK and not just to more deprived areas of the UK such as Salford. To the best of our knowledge, no study has reported the impact of the COVID-19 pandemic on HbA_1c_ monitoring in diabetes, and no study has described national variation in mortality rates in people with type 2 diabetes following the first peak of the pandemic.

Our data have important clinical implications. In early March 2020, GPs were advised to minimise the number of face-to-face contacts they had with their patients, including NHS health-checks ^12^. Our data suggests that this reduction of clinical services has led to major reductions in the diagnosis and monitoring of type 2 diabetes. Whilst the decline in new metformin prescriptions was less evident than the reduction in the rate of new diabetes diagnoses (53% vs. 70%), which is potentially due to people with prevalent diabetes receiving treatment intensification with metformin, these concomitant reductions in new prescriptions issued for metformin and insulin further support these findings.

Type 2 diabetes develops over many years, so it seems unlikely that people’s behaviour during the pandemic has reduced the true incidence of these conditions. If we assume that the true incidence of type 2 diabetes has remained constant from March 2020, our data suggest that, across the UK, the indirect consequences of the pandemic have led to nearly 60K missed/delayed diagnoses of type 2 diabetes in the nine months between 1^st^ March and 10^th^ December 2020 (**Supplementary Table S3**). This figure may be an underestimate if changes in lifestyle during lockdown have increased obesity rates or other risk factors for diabetes in the general population^13^. In cross-sectional surveys of UK adults conducted during the first lockdown in the UK (April-May 2020), participants reported fewer behaviours protecting weight gain compared to before lockdown. This included aspects of diet, physical activity, alcohol consumption, self-perceived mental health and sleep quality ^14–18^. Lockdown may also have had a disproportionally larger influence on weight-related behaviours among those with higher BMI ^14^. During lockdown, higher BMI was associated with lower levels of physical activity and diet quality, and a greater frequency of overeating relative to people with lower BMI ^14^. These data are a clinical concern because undiagnosed type 2 diabetes could contribute to serious long-term complications ^5^.

The huge reduction in the rate of HbA_1c_ testing is another important concern for people with type 2 diabetes, because they, and their clinicians, often rely solely on HbA_1c_ data to make treatment decisions. The reduction in new prescriptions for insulin was largely observed in older individuals suggesting this reduction was explained by a failure to intensify therapy in people with poorly controlled long-duration type 2 diabetes. There are already concerns in the UK about clinical inertia in diabetes management, with frequent failures to escalate care when glucose control is poor ^19^. These HbA_1c_ data indicate potential further delays in the management of type 2 diabetes that are predicted to cause avoidable diabetes-related long-term complications. A reduced frequency of HbA_1c_ testing in primary care might also contribute to missing people with non-diabetic hyperglycaemia who might benefit from referral to the NHS Diabetes Prevention Programme.

The higher COVID-related death rate in people with diabetes has been well-documented ^2,3,20^, and our data support these observations. Here, we add to these data by showing national differences in the impact of COVID-19 on mortality rates in people with type 2 diabetes, with higher rates observed in England compared to the rest of the UK. Further research is required to understand how population characteristics including ethnicity, population density and deprivation might explain these differences.

As engagement with health services increases, and hopefully is maintained during subsequent COVID-19 peaks, our data predict a marked increase in presentations with incident type 2 diabetes. Should this occur, then healthcare services will need to manage this backlog, and the anticipated greater deterioration of diabetes parameters including HbA_1c_ brought about by delayed diagnoses. Older individuals, males and people from deprived backgrounds appear to be most adversely affected by reductions in rates of diagnosis and monitoring of type 2 diabetes. As outpatient diabetes services start to open up, these individuals may be a group to target for early intervention, and in particular, for HbA_1c_ testing and treatment intensification when appropriate. During this pandemic and its associated lockdowns, effective public communications should ensure that patients remain engaged with diabetes services including HbA_1c_ screening and monitoring ^21^, and make use of remote consultations ^22,23^.

Our study had several strengths: this is the first UK-wide study reporting the indirect impact of the COVID-19 pandemic on the diagnosis of type 2 diabetes, related prescribing and HbA_1c_ testing in primary care. Our findings in English practices were replicated using data from other parts of the UK. By combining assessments of diabetes coding and prescribing, our data supports the conclusion that reduced rates of diagnoses are genuinely explained by missed diagnoses. Our study has some limitations: First, ethnicity coding is not adequately captured in primary care and therefore we had limited ability to explore ethnicity-related variation in care and outcomes. Future studies will incorporate linked secondary care data that has more complete capture of ethnicity data. Second, it is possible that some diabetes diagnoses may have been made in a hospital setting following an acute presentation and that the related primary care coding had not been updated at the time of our data extraction. While hospital presentation of incident diabetes may have occurred in some instances, it would not explain the reductions in new prescribing for metformin and this potential explanation does not fit with our local experience. In general, people have avoided hospital attendance during the pandemic. For example, one study documented a 23% reduction in emergency admissions in the UK ^24^. Finally, although our results and conclusions are relevant to the UK population, generalisability to other healthcare systems may be limited. However, a pan-European study of diabetes specialist nurses reported that diabetes services and the level of care provided to people with diabetes had been significantly disrupted during the pandemic with perceived reductions in new diabetes diagnoses, routine care provision, diabetes education and psychological support ^25^.

In conclusion, we highlight marked reductions in the diagnosis and monitoring of type 2 diabetes as indirect consequences of the COVID-19 pandemic. Over the coming months, healthcare services will need to manage this predicted backlog, and the anticipated greater deterioration of diabetes parameters due to delayed diagnoses and reduced monitoring in those with established diabetes. Older people, men and those from deprived backgrounds with type 2 diabetes will be specific groups to target for early HbA_1c_ testing and intervention. During this pandemic and associated lockdowns, effective public communications should ensure that patients remain engaged with diabetes services including HbA_1c_ screening and monitoring and make use of remote consultations.

## Supporting information

RECORD checklist

## Data Availability

The clinical codes used in this study are published on ClinicalCodes.org. Electronic health records are, by definition, considered "sensitive" data in the UK by the Data Protection Act and cannot be shared via public deposition because of information governance restriction in place to protect patient confidentiality. Access to data are available only once approval has been obtained through the individual constituent entities controlling access to the data. The primary care data can be requested via application to the Clinical Practice Research Datalink.

https://www.cprd.com/

https://clinicalcodes.rss.mhs.man.ac.uk/

## ACKNOWLEDGEMENTS

The study is based on data from the Clinical Practice Research Datalink obtained under license from the UK Medicines and Healthcare products Regulatory Agency. The study and use of CPRD data was approved by the Independent Scientific Advisory Committee for Clinical Practice Research Datalink research (protocol number: 20_182R). The data is provided by patients and collected by the NHS as part of their care and support. The interpretation and conclusions contained in this study are those of the authors alone. We would like to acknowledge all the data providers and general practices that made the anonymised data available for research.

## Data availability

All clinical codes used in the study are published on Clinicalcodes.org. Electronic health records are, by definition, considered “sensitive” data in the UK by the Data Protection Act and cannot be shared via public deposition because of information governance restriction in place to protect patient confidentiality. Access to data are available only once approval has been obtained through the individual constituent entities controlling access to the data. The primary care data can be requested via application to the Clinical Practice Research Datalink (www.cprd.com).

## Funding

This work is funded by the National Institute for Health Research (NIHR) Greater Manchester Patient Safety Translational Research Centre. The views expressed are those of the authors and not necessarily those of the NIHR or the Department of Health and Social Care.

The funding source had no role in the study design, data collection, data analysis, data interpretation, or writing of the report.

## Authors’ relationships and activities

All authors have completed the Unified Competing Interest form (available on request from the corresponding author) and declare: no support from any organisation for the submitted work; no financial relationships with any organisations that might have an interest in the submitted work in the previous three years. DMA reports research funding from AbbVie, Almirall, Celgene, Eli Lilly, Novartis, Janssen, UCB and the Leo Foundation outside the submitted work. MKR has received consulting fees and non-promotional lecture fees from Novo Nordisk in relation to cardiovascular disease and diabetes. The company has had no role in influencing the proposed study and is not expected to benefit from this work. Outside the submitted work, MKR reports receiving research funding from Novo Nordisk, consultancy fees from Novo Nordisk and Roche Diabetes Care, and modest owning of shares in GlaxoSmithKline. NM reports honorarium for presentations from Napp Pharmaceuticals, Novo Nordisk, Sanofi, MyLan, Boehringer Ingelheim, Lilly Diabetes, Abbott, Omnia-Med, Takeda UK and AstraZeneca. All other authors declare no competing interests. There are no other relationships or activities that could appear to have influenced the submitted work.

## Contribution statement

DMA conceived the original idea. MKR, MJC and AKW helped develop the idea. MJC and AKW performed the analysis and verified the analytical methods. MKR and DMA also reviewed the clinical code sets. NK and NM (primary care clinicians) and LL, HT (secondary care diabetes clinicians) helped interpret the results. MKR wrote the manuscript with input from all authors. All authors critically reviewed and approved the final version.

The lead authors (MJC and AKW: the manuscript’s guarantors) affirm that the manuscript is an honest, accurate, and transparent account of the study being reported; that no important aspects of the study have been omitted; and that any discrepancies from the study as planned have been explained. The corresponding author had full access to all of the data and the final responsibility to submit for publication.

## Supplementary Material

**Supplementary Figure S1.**
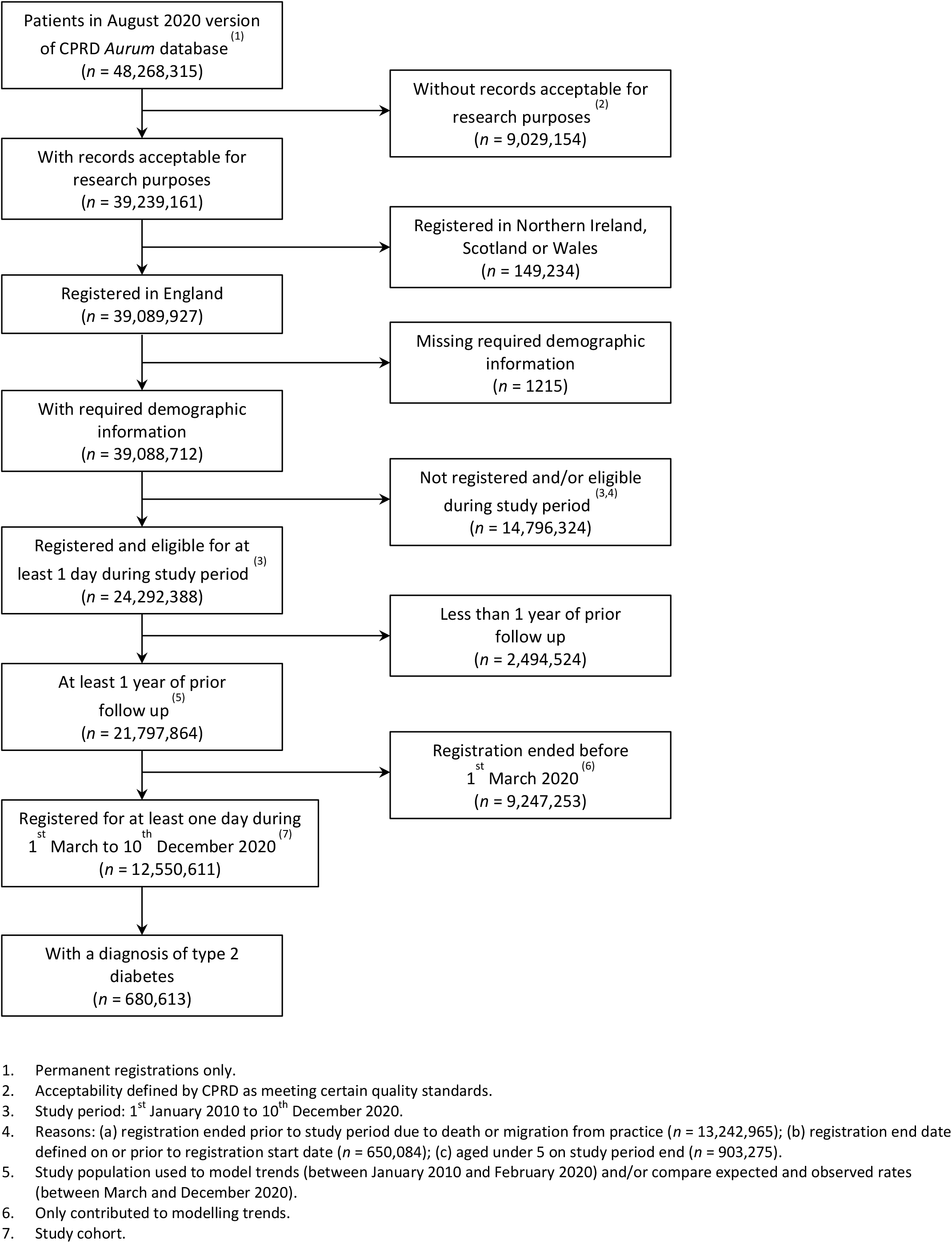
Flow diagram of the cohort from CPRD *Aurum*

**Supplementary Figure S2.**
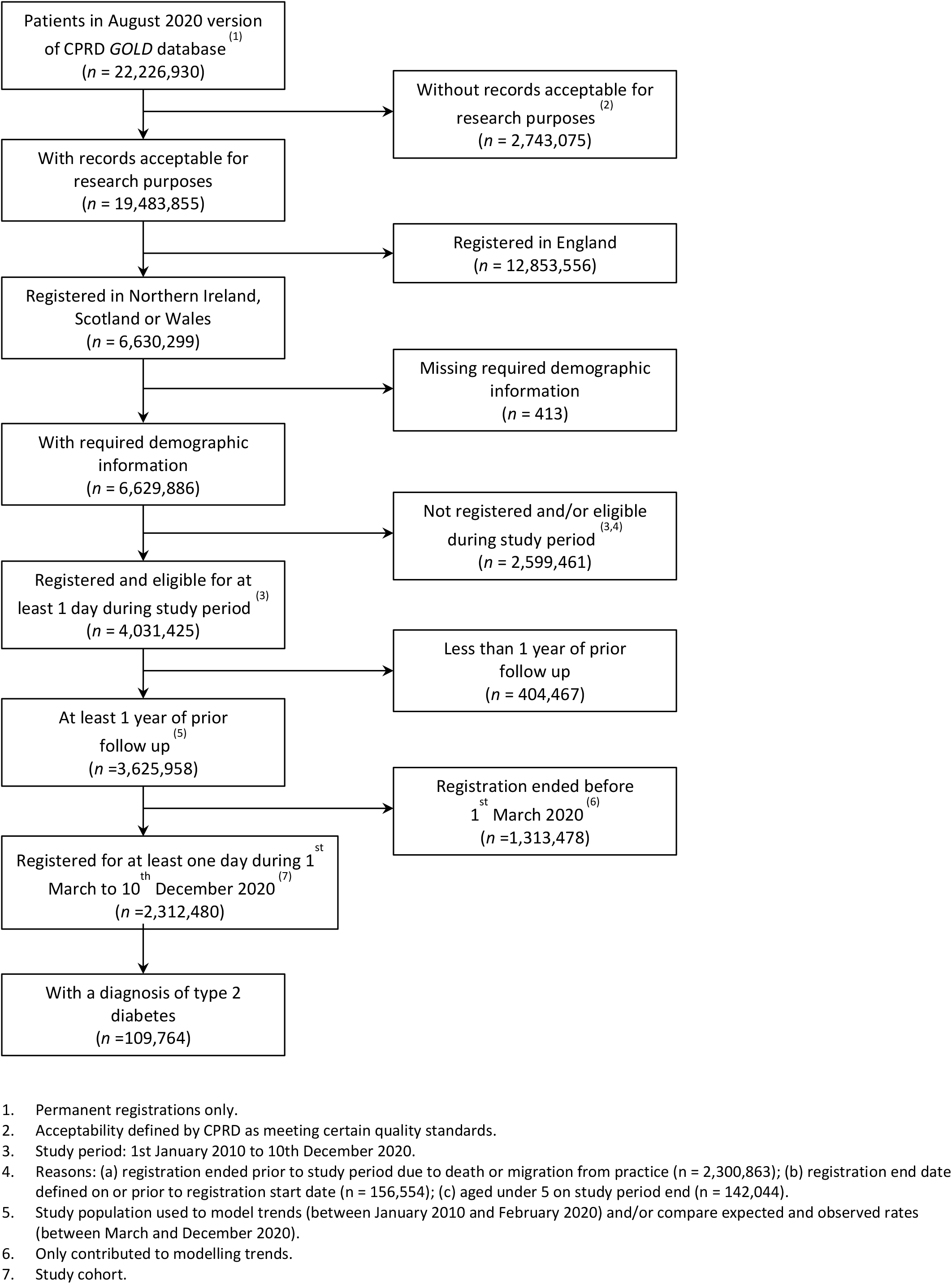
Flow diagram of the cohort from CPRD *GOLD*

**Supplementary Table S1.**
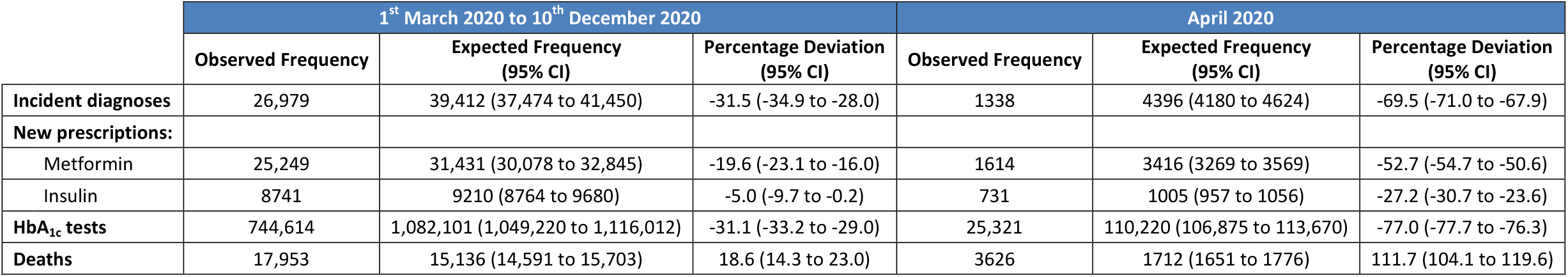
Comparison of observed and expected monthly incidence rates for type 2 diabetes in primary care, HbA_1c_ monitoring in type 2 diabetes, new prescriptions for metformin and insulin, and deaths in people with type 2 diabetes between 1^st^ March 2020 and 10^th^ December 2020 and during the first COVID-19 peak in April 2020, in England (CPRD *Aurum*).

**Supplementary Figure S3.**
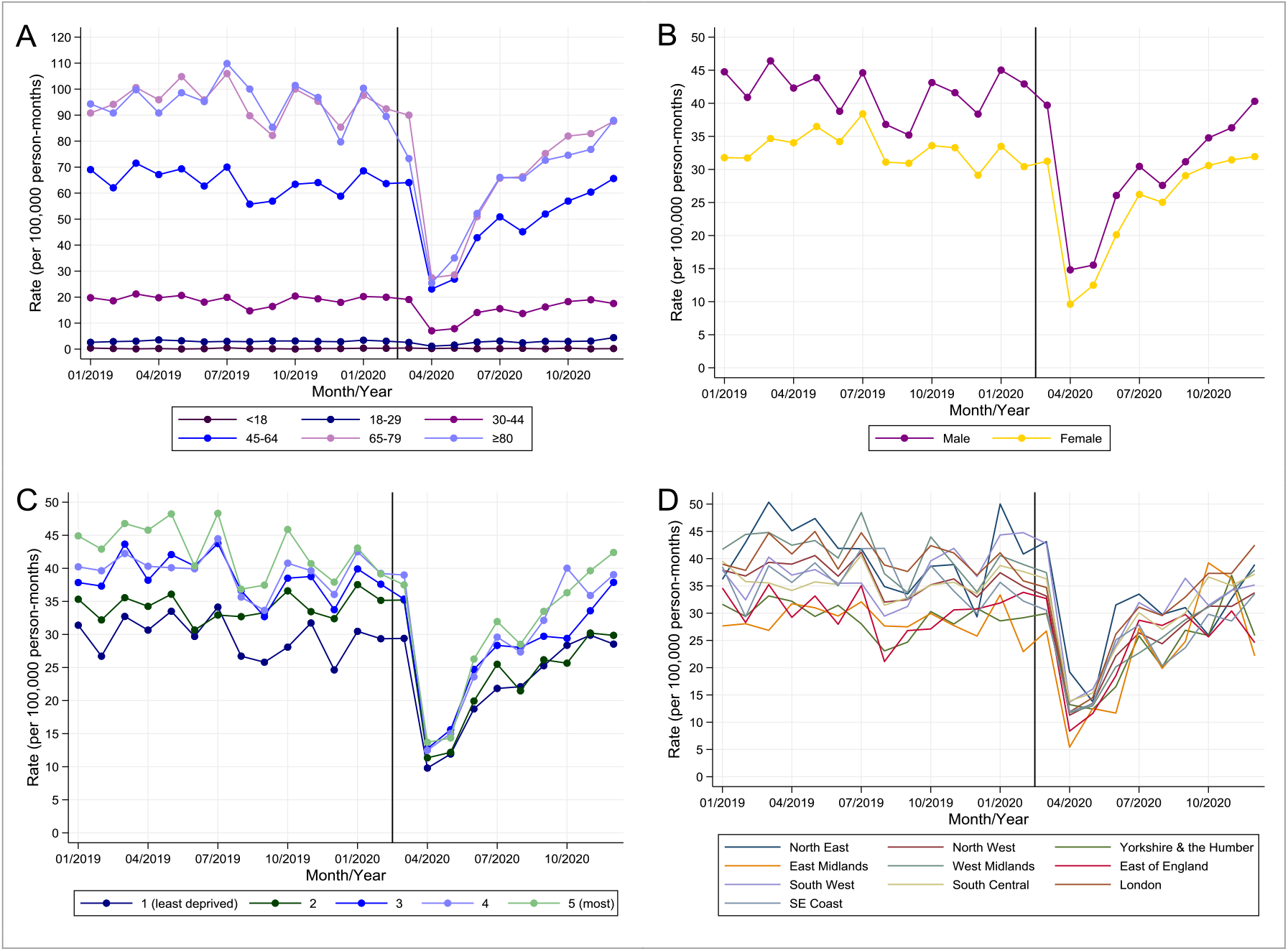
Comparison of monthly incidence rates for type 2 diabetes in primary care before and after the start of the COVID-19 pandemic in England (CPRD *Aurum*). Rates are stratified by age group (*A*), gender (*B*), Index of Multiple Deprivation quintile (*C*), and by region (*D*).

**Supplementary Figure S4.**
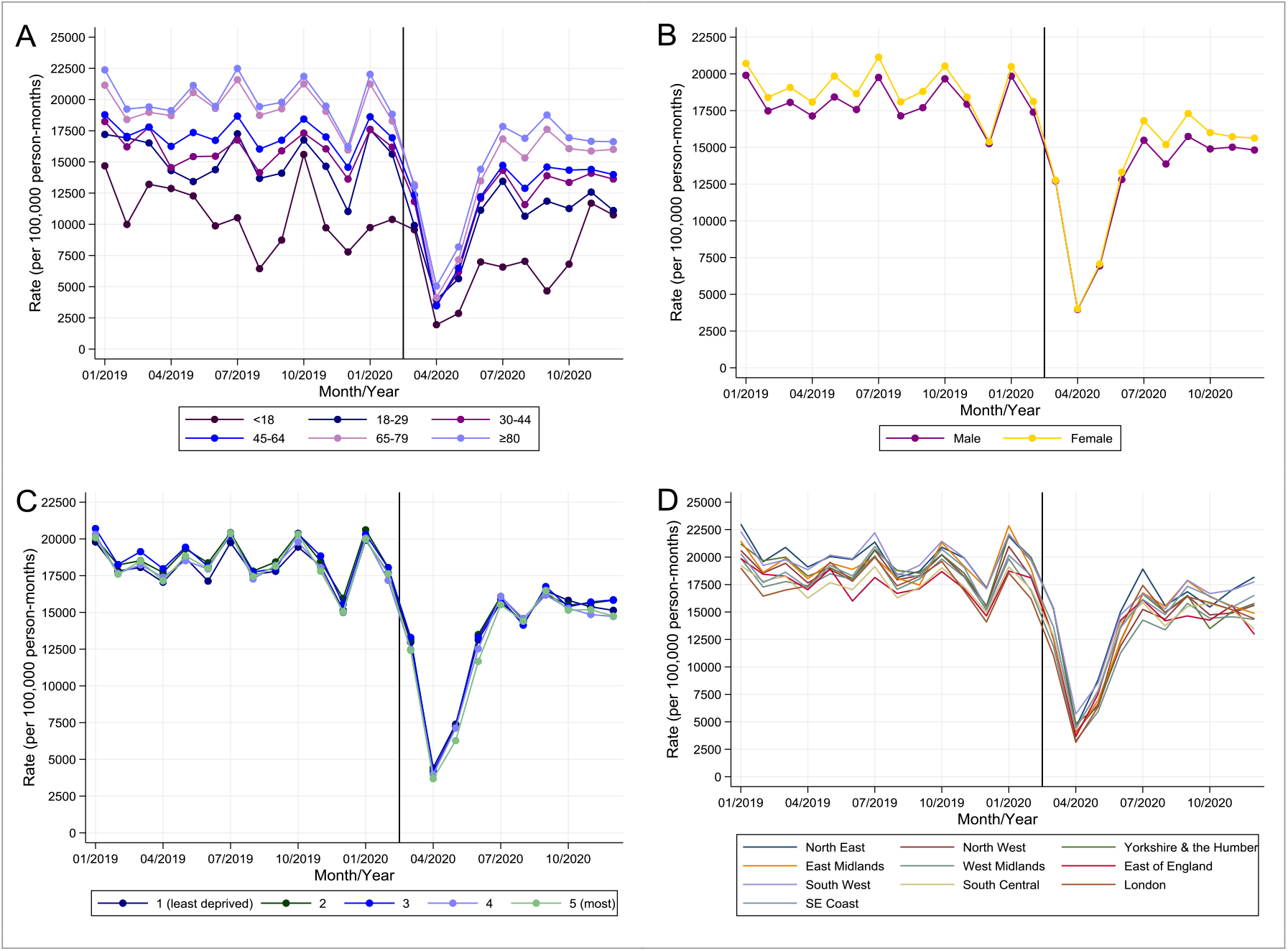
Comparison of monthly HbA_1c_ testing rates in people with type 2 diabetes in primary care before and after the start of the COVID-19 pandemic in England (CPRD *Aurum*). Rates are stratified by age group (*A*), gender (*B*), Index of Multiple Deprivation quintile (*C*), and by region (*D*).

**Supplementary Figure S5.**
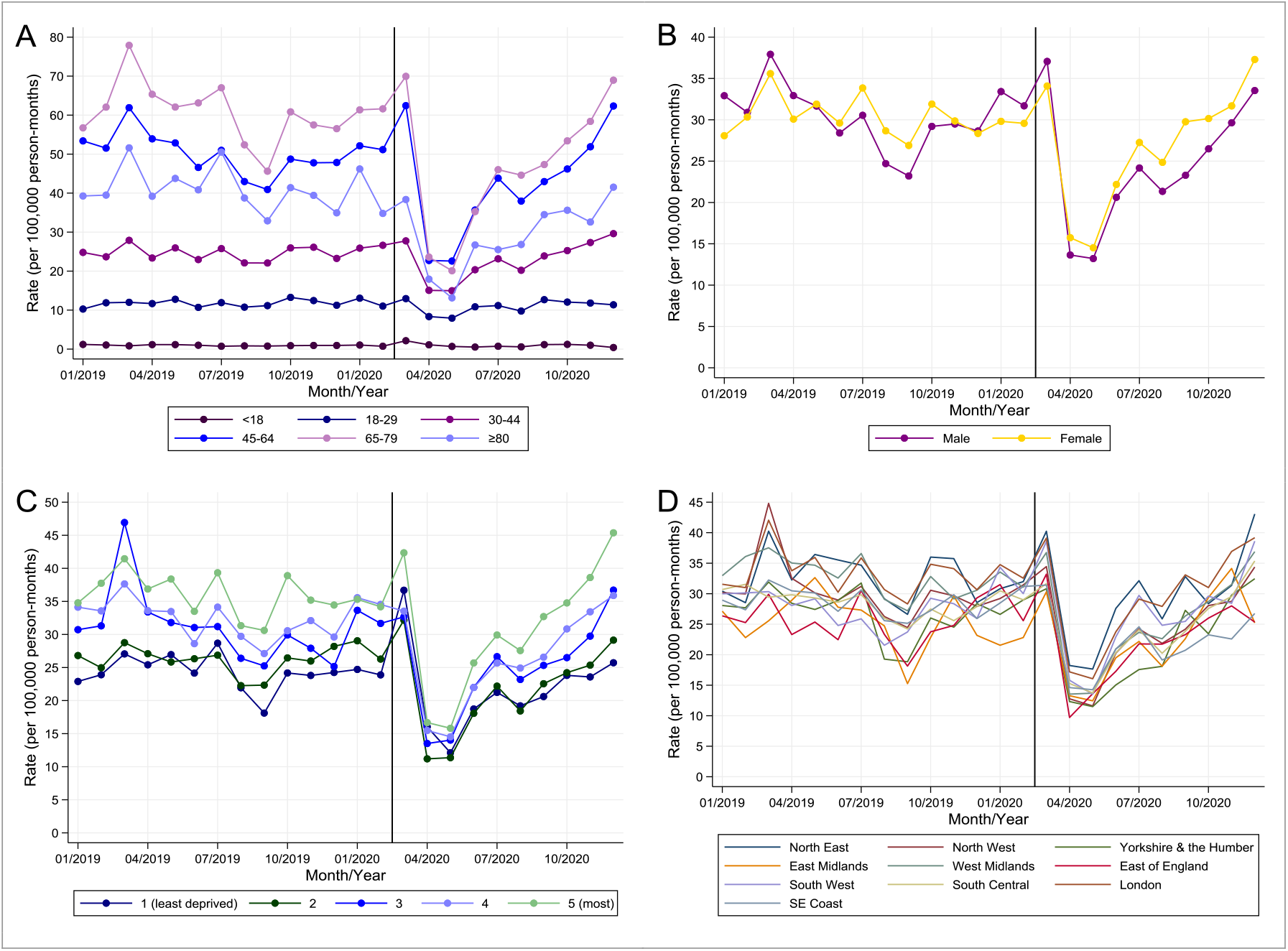
Comparison of monthly incidence rates for metformin prescribing in primary care before and after the start of the COVID-19 pandemic in England (CPRD *Aurum*). Rates are stratified by age group (*A*), gender (*B*), Index of Multiple Deprivation quintile (*C*), and by region (*D*).

**Supplementary Figure S6.**
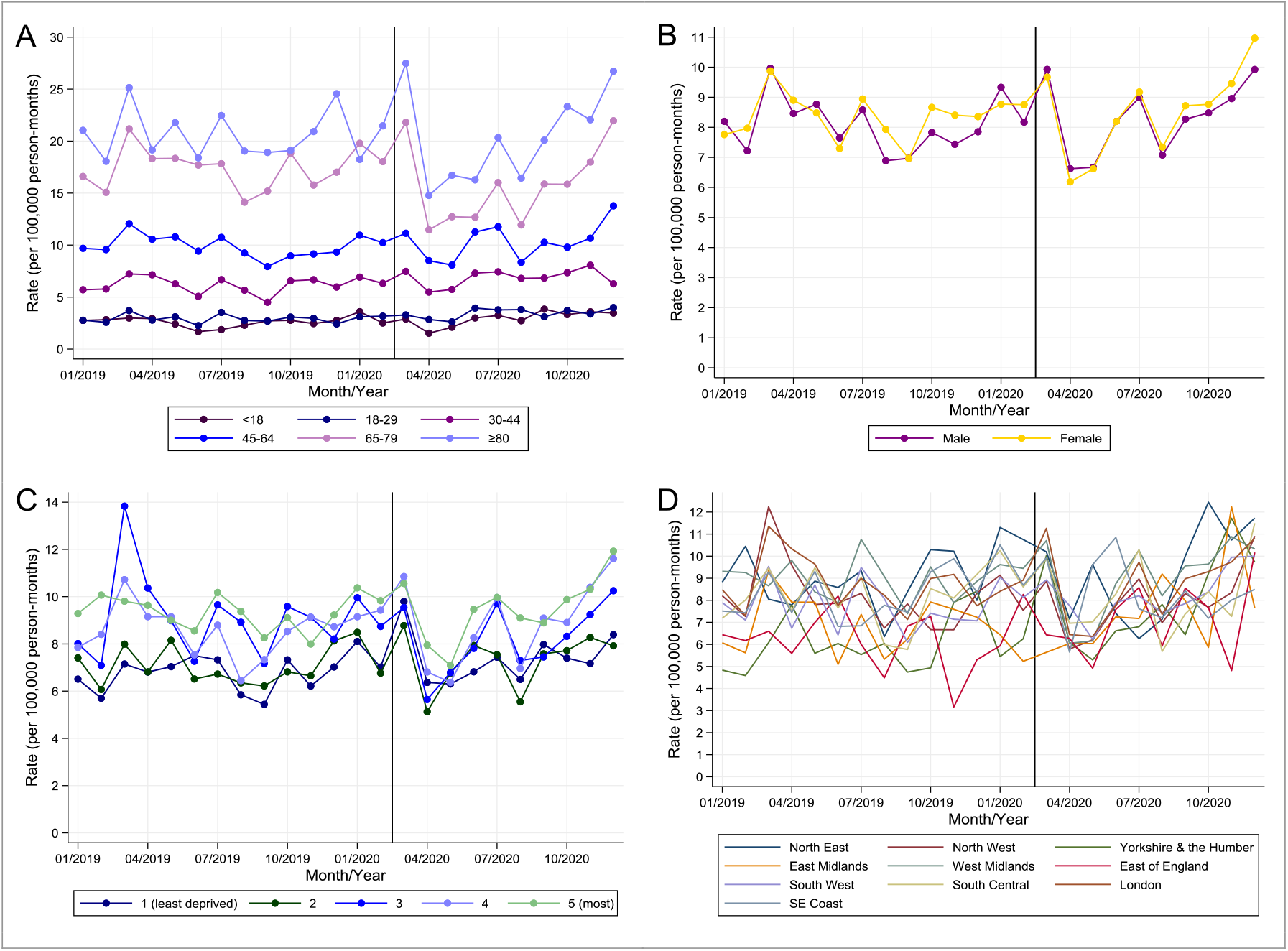
Comparison of monthly incidence rates for insulin prescribing in primary care before and after the start of the COVID-19 pandemic in England (CPRD *Aurum*). Rates are stratified by age group (*A*), gender (*B*), Index of Multiple Deprivation quintile (*C*), and by region (*D*).

**Supplementary Figure S7.**
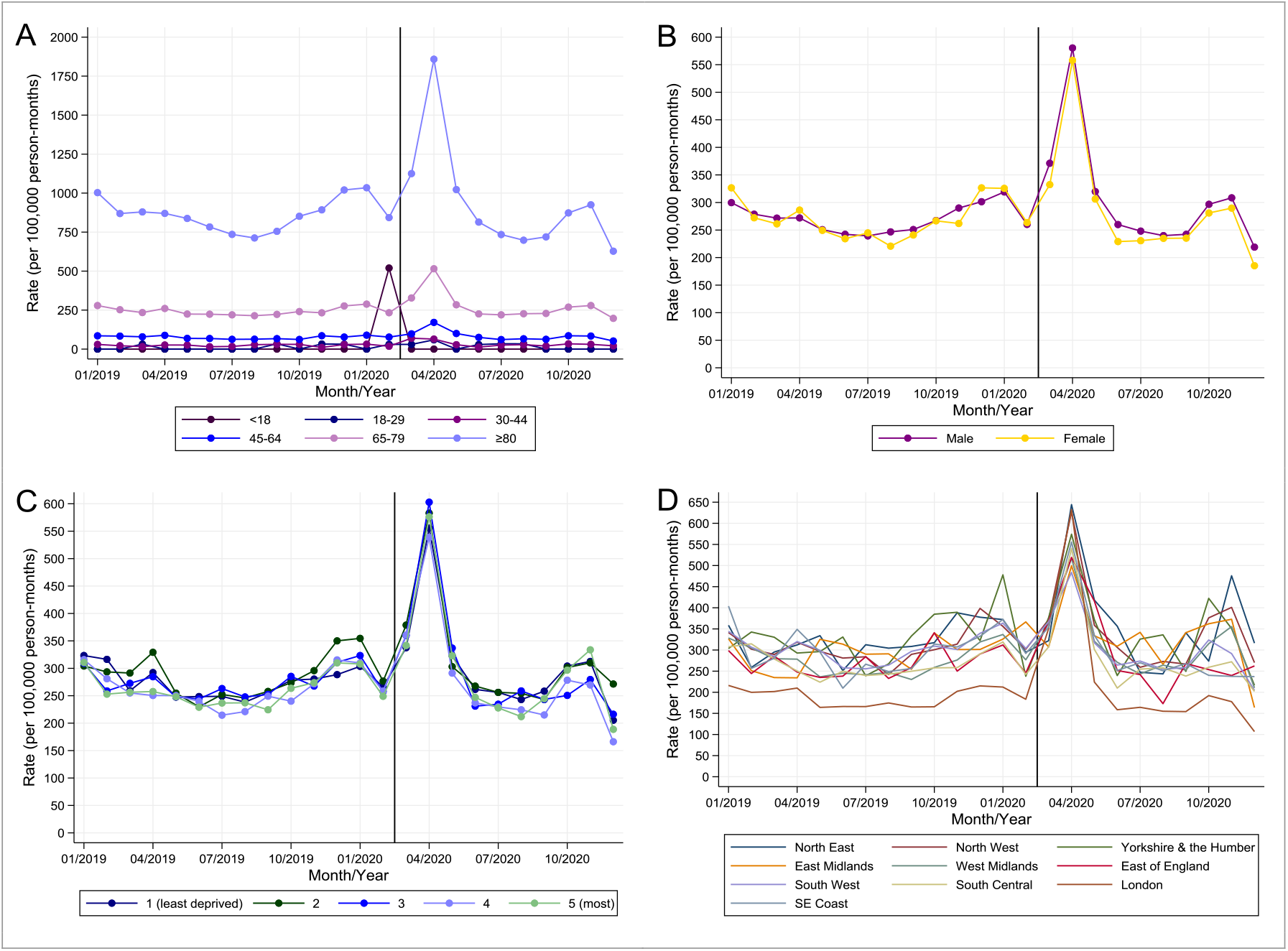
Comparison of monthly mortality rates in people with type 2 diabetes in primary care before and after the start of the COVID-19 pandemic in England (CPRD *Aurum*). Rates are stratified by age group (*A*), gender (*B*), Index of Multiple Deprivation quintile (*C*), and by region (*D*).

**Supplementary Table S2.** Comparison of observed and expected monthly incidence rates for type 2 diabetes in primary care, HbA_1c_ monitoring in type 2 diabetes, new prescriptions for metformin and insulin, and deaths in people with type 2 diabetes between 1^st^ March 2020 and 10^th^ December 2020 and during the first COVID-19 peak in April 2020, in Northern Ireland, Scotland and Wales (CPRD *GOLD*).

|  | 1<sup>st</sup> March 2020 to 10<sup>th</sup> December 2020 |  |  | April 2020 |  |  |
| --- | --- | --- | --- | --- | --- | --- |
|  | Observed Frequency | Expected Frequency<br>(95% CI) | Percentage Deviation<br>(95% CI) | Observed Frequency | Expected Frequency<br>(95% CI) | Percentage Deviation<br>(95% CI) |
| **Incident diagnoses** | 4865 | 6190 (5823 to 6581) | -21.4 (-26.0 to -16.4) | 221 | 701 (659 to 745) | -68.4 (-70.3 to -66.4) |
| **New prescriptions:** |  |  |  |  |  |  |
| Metformin | 5019 | 5672 (5365 to 5996) | -11.5 (-16.2 to -6.4) | 266 | 629 (595 to 665) | -57.7 (-60.0 to -55.2) |
| Insulin | 1607 | 1712 (1605 to 1825) | -6.1 (-11.9 to 0.1) | 148 | 188 (177 to 201) | -21.2 (-26.3 to -16.3) |
| **HbA<sub>1c</sub> tests** | 84,412 | 133,011 (128,805 to 137,354) | -36.5 (-38.5 to -34.4) | 2245 | 13,455 (13,030 to 13,894) | -83.3 (-83.8 to -82.7) |
| **Deaths** | 3145 | 2773 (2650 to 2900) | 13.4 (8.4 to 18.6) | 504 | 305 (291 to 319) | 65.2 (57.9 to 73.1) |

**Supplementary Figure S8.**
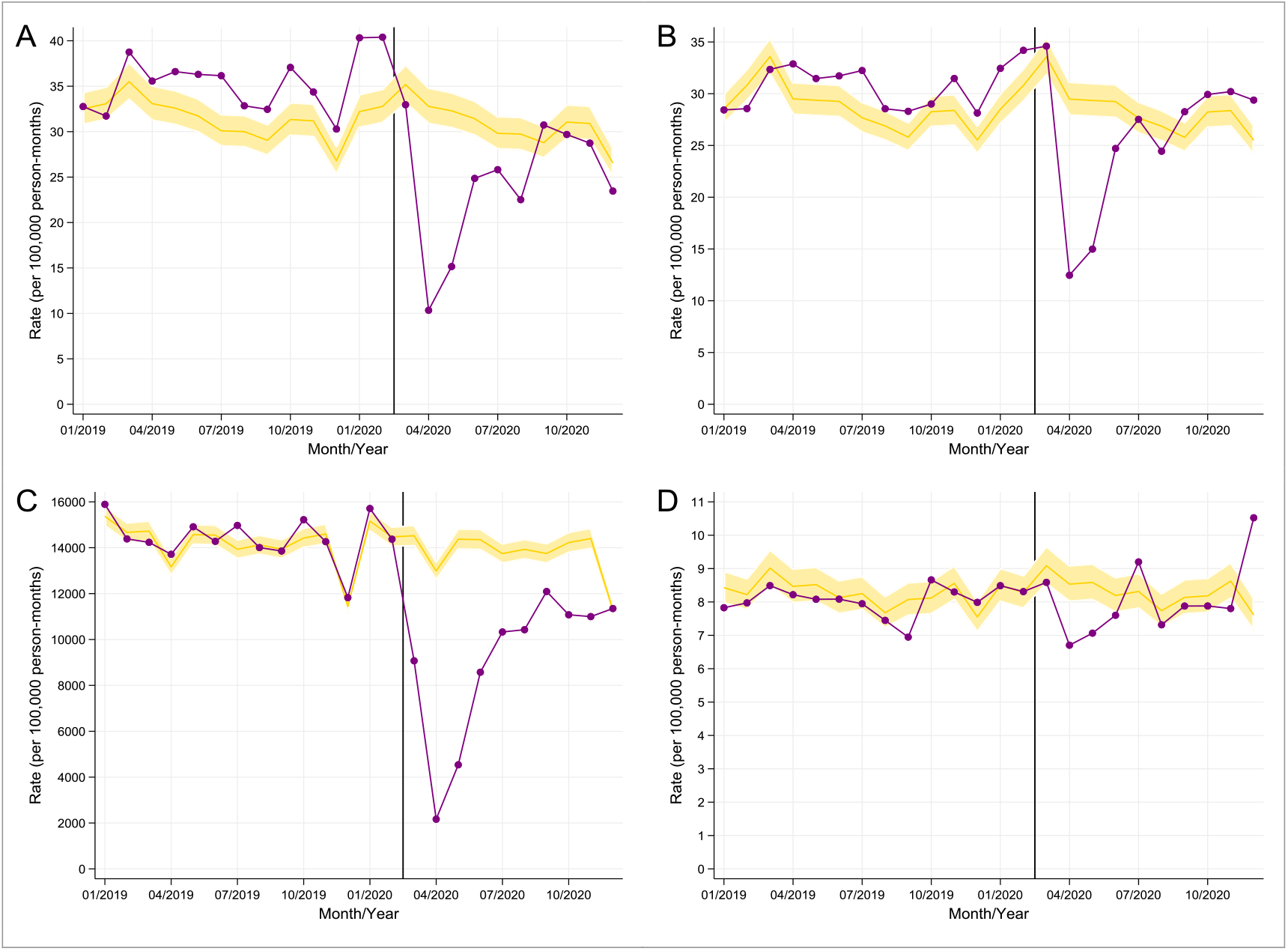
Comparison of observed (purple line) and expected (gold line with shaded area representing 95% CI) monthly rates in primary care before and after the start of the COVID-19 pandemic in Northern Ireland, Scotland, and Wales. *A*: incident diagnoses of type 2 diabetes, *B*: new metformin prescriptions, *C*: HbA_1c_ monitoring in patients with type 2 diabetes, *D*: new insulin prescriptions. Rates were derived using data from CPRD *GOLD* covering 3,625,958 patients. X-axis markers are mid-month. The vertical line denotes 1^st^ March 2020.

**Supplementary Table S3.** Estimating the number of missed or delayed diagnoses of type 2 diabetes in the UK population between March and December 2020.

|  | Population * |  | Sample |  | Estimate of missed/delayed T2DM diagnoses |  |
| --- | --- | --- | --- | --- | --- | --- |
|  | All ages | Age>=5 | Age>=5 | % | Sample | Population |
| **UNITED KINGDOM** | 66,796,807 | 62,939,544 | 14,863,091 | 23.61 |  |  |
| **ENGLAND** | 56,286,961 | 52,987,324 | 12,550,611 | 23.69 | 12,433 | 52,491 |
| **NI, SCOTLAND, WALES** | 10,509,846 | 9,952,220 | 2,312,480 | 23.24 | 1325 | 5702 |
|  |  |  |  |  |  | 58,193 |

\* Source: https://www.ons.gov.uk/peoplepopulationandcommunity/populationandmigration/populationestimates

## Notes

### Funding Statement

This work was funded by the National Institute for Health Research (NIHR) Greater Manchester Patient Safety Translational Research Centre. The views expressed are those of the authors and not necessarily those of the NIHR or the Department of Health and Social Care. The funders of the study had no role in study design, data collection, data analysis, data interpretation, or writing of the report.

### Author Declarations

This study is based on data from the Clinical Practice Research Datalink obtained under license from the UK Medicines and Healthcare products Regulatory Agency. The study and use of CPRD data was approved by the Independent Scientific Advisory Committee for Clinical Practice Research Datalink research (protocol number: 20_182R).

### Summary of Updates

The original submission has now been updated using an extension of the data to December 2020. All tables, figures and supplemental files have been updated accordingly.

